# Circulating ADAM17 is associated with COVID-19 severity

**DOI:** 10.1101/2023.08.23.23294465

**Authors:** Mengyu Pan, Isabel Goncalves, Andreas Edsfeldt, Jiangming Sun, Per Swärd

## Abstract

**Background:** ADAM17 are emerging as an important role in the severe outcomes of COVID-19. This study aims to characterize causal relationship between ADAM17 and COVID-19.

**Methods:** Using mendelian randomization analyses, we examined the causal effects for circulating ADAM17 on COVID-19 outcomes using summary statistics from large genome wide association studies of ADAM17 (up to 35 559 individuals) from the Icelandic Cancer Project and deCODE genetics, critical COVID-19 (cases:13 769; controls:1 072 442), hospitalized COVID-19 (cases:32 519; controls: 2 062 805) and SARS-CoV-2 infection (cases:122 616; controls:2 475 240) from the COVID-19 Host Genetics Initiative.

**Results:** Mendelian randomization analyses demonstrated that 1 standard deviation increase of genetically determined circulating ADAM17 at extracellular domain were associated with increasing risk of developing critical COVID-19 (odds ratio [OR]=1.26, 95% CI 1.03-1.55). Multivariable MR analysis suggested a direct causal role of circulating ADAM17 at extracellular domain on the risk of critical COVID-19 (OR=1.09; 95% CI 1.01-1.17), accounting for body mass index. Casual effects for the cytoplasmic domain of ADAM17 on COVID-19 were not observed.

**Conclusion:** Our results suggest that the increased circulating ADAM17 at extracellular domain are associated with a high risk of critical COVID-19 strengthening that of ADAM17 may contribute to the risk stratification and a therapeutic option for severe COVID-19.

**What is already known on this topic:** Various inflammatory stimuli, as well as the SARS-CoV-2 S-protein, elevate the activity of a disintegrin and metalloproteinase 17 (ADAM17). Inhibition of ADAM17 activity *in vitro* has illustrated the ability to effectively impede the infection caused by SARS-CoV-2. Nonetheless, the predictive capability of ADAM17 in predicting the severity of COVID-19 outcomes remains less certain within human populations.

**What this study adds:** Using large genome wide association studies, Mendelian randomization study demonstrated that genetic susceptibility to the increased circulating levels of ADAM17 (extracellular domain) were associated with critical COVID-19 but not SARS-CoV-2 infection or hospitalized COVID-19.

**How this study might affect research, practice or policy:** The study’s insights might pave the way for novel therapeutic strategies targeting ADAM17 activity. Patients with a genetic predisposition to higher ADAM17 activity might be identified and given tailored treatments.

## Introduction

Infection with severe acute respiratory syndrome coronavirus 2 (SARS-CoV-2) can range from asymptomatic to severe with risk of acute respiratory distress syndrome (ARDS), multiorgan failure and death (1). The entrance of SARS-COV-2 in host cells is made possible by the binding of the viral S-protein to the active surface domain of angiotensin converting enzyme 2 (ACE2) (2). Normally, ACE2 exerts cardiovascular protective effects by cleaving angiotensin (ANG) II into ANG 1-7 which has vasodilatory, anti-inflammatory and anti-fibrotic effects. Various inflammatory stimuli and the SARS-COV-2 S-protein increase a disintegrin and metalloproteinase 17 (ADAM17) activity, leading to the shedding of the catalytically active ectodomain of ACE2 (3, 4). ADAM17 gene expression is normally high in the lung (www.gtxportal.com), and increased ADAM17 activity could in addition to shedding of ACE2 also play a pivotal role to induce COVID-19-associated lung inflammation by shedding membrane-bound tumour necrosis factor (TNF) alpha, interleukin (IL)6R, TNF receptors (TNFRs) and other pro-inflammatory mediators, contributing to the cytokine storm observed in severe COVD-19 (5). Overactivation of ADAM17 activity may be of particular importance in COVID-19, given the direct activation of this protease by the virus. Also, the inhibition of ADAM17 activity in vitro has been demonstrated to effectively counteract SARS-CoV-2 infection. Collectively, these findings underscore the growing significance of ADAM17 as a pivotal mediator in the context of severe COVID-19 (6-8). However, the affirmation of this statement through population studies remains constrained, as there is limited evidence indicating whether ADAM17 levels can serve as prognostic indicators for severe COVID-19 outcome.

Circulating ADAM17 retains its proteolytic activity, and may reflect tissue-bound ADAM17 (8). Linking high levels of ADAM17 to increased risk of severe COVID-19 would strengthen that ADAM17 inhibition could be a promising therapeutic option against COVID-19. The regulation of membrane-bound ADAM17 is highly complex. At the post-translational level, interaction with native inhibitors, native activators, adapter proteins and phosphorylation status are essential for the transport to the cell surface (5). Of particular importance, iRhom2 upregulation is important for ADAM17 activity under pro-inflammatory conditions (3). Therefore, to answer the query of ADAM17’s potential role in severe COVID-19 in humans, it is more relevant to assess levels of membrane-bound ADAM17, rather than the gene expression of ADAM17. It is also important to discriminate between the cytoplasmic and extracellular domains given that the cytoplasmic domain may be hidden from detection in plasma samples (8). Therefore, by employing mendelian randomization we aimed to investigate if there is a causal relationship between circulating ADAM17 (cytoplasmic and membrane-bound domains) and the risk of severe COVID-19 using the latest genome-wide association studies (GWAS) of ADAM17 (9) and COVID-19 (10).

## Methods

### Data sources

Summary statistics for GWAS of ADAM17 were retrieved from large GWAS of plasma protein levels measured in 35 559 middle-aged European populations (mean age=55 years old, standard deviation=17 years, 57% were women) (9). Protein levels in plasma samples were measured with the SomaScan aptamers (SomaLogic, Inc. USA), and two different aptamers were used for ADAM17; one which measures the extracellular and another the cytoplasmic domain. Protein levels were rank-inverse normal transformed adjusting for age, sex and sample age in GWAS analysis of ADAM17.

GWAS summary statistics for COVID were retrieved from the COVID-19 Host Genetics Initiative (release 7) (10). As reported, GWAS of COVID-19 disease was performed using cases defined by: (1) critically ill cases of COVID-19 (cases:13 769; controls:1 072 442) who required respiratory support in hospital or who died due to the disease, (2) moderate cases of COVID-19 who were hospitalized (cases:32 519; controls: 2 062 805), and (3) all reported SARS-CoV-2 infected cases (cases:122 616; controls:2 475 240). Controls were genetically ancestry-matched samples without previous known SARS-CoV-2 infection.

GWAS of body-mass index (BMI, n=806 834) were retrieved from a GIANT and UK BioBank Meta-analysis (11) for multivariable Mendelian randomization (MR) analysis.

GWAS restricted to European population were used in the present MR analyses. Genomic positions for single nucleotide polymorphism (SNPs) were harmonized to the same strand in human genome build 19. Ambiguous SNPs and SNPs with a non-inferable forward strand were excluded. Palindromic SNPs with difference in effect allele frequencies greater than 0.2 were also excluded.

Details on publicly available GWAS summary statistics were provided in the Supplemental Table 1 in the Supplemental Material.

### Instrumental Variable Selection

If not specific, independent variants were clumping with (linkage disequilibrium (LD) r^2^ < 0.001 in a window of 500 kb) showed genome-wide significance (*P* < 5 × 10^−8^) in respective exposure (ADAM17 or COVID-19) were chosen as instruments. The p-value for instrumental variants-outcome association is greater than 1 x 10^−5^. When the exposure is ADAM17 (cytoplasmic domain), *P* < 1 × 10^−6^ were used as a cutoff to select instruments since as quite a smaller number of SNPs were genome-wide associate with ADAM17 (cytoplasmic domain) levels. Average F-statistic for the selected instrumental variable was reported to present its strength for respective exposure.

### Primary Mendelian randomization analyses

MR were implemented using the inverse-variance weighted (IVW) to examine causal effects for circulating ADAM17 (cytoplasmic domain and cytoplasmic domain, respectively) on COVID-19 (Critical, hospitalized and SARS-CoV-2 infected). Reversely, causal effects for COVID-19 on circulating ADAM17 were also tested using IVW. Proxy variants were not used if variants were not available in the out-come GWAS. Heterogeneity in effect size between instrumental variants was assessed by Cochran’s Q statistic. Potential horizontal pleiotropy was examined by MR-Egger (12).

### Sensitivity MR analyses

Using same settings of primary MR analysis, we conducted sensitivity analyses by weighted median (13) and MR-Egger (14). The weighted median method assumes that most genetic variants are valid instrumental variables which is robust to outliers but sensitive to additional/removal of SNPs into instrumental variables. MR-Egger can test for directional pleiotropy and estimate causal effect under InSIDE (INstrument Strength Independent of Direct Effect) assumption whereas InSIDE is often not plausible suggesting that MR-Egger may be less efficient (14).

Additional analyses were also performed using robust adjusted profile score (RAPS) (15) by setting cut-off of p-value of 0.001 where other settings same as primary MR analysis. MR-RAPS which can account for weak instrument bias, extreme outliers and pleiotropic effects of SNPs by assuming pleiotropic effects are normally distributed. MR-RAPS performed well when the assumption is fulfilled but not when violated.

### Multivariable MR analysis

Multivariable MR analysis was implemented on genetic variants that were associate with either BMI or ADAM17 (*P* < 5 × 10^−8^) using IVW method. Variants were pruned to exclude associated variants with LD r^2^ greater than 0.001.

MR analyses were implemented using the R (version 4.1.3) package TwoSampleMR (version 0.5.6) (16).

## Results

GWAS of ADAM17 recruited 35,559 Icelanders where 52% of participants were from the Icelandic Cancer Project and 48% deCODE. GWAS of the hospitalized COVID-19 and SARS-CoV-2 infection also employed 89 cases and 274 322 controls from deCODE. GWAS of critical COVID-19 didn’t include deCODE. Hypothetically, the largest possible overlapped samples accounted for 48% of total subjects in GWAS of ADAM17, 13% for GWAS of the hospitalized COVID-19, 11% for GWAS of SARS-CoV-2 infection and 0% for GWAS of critical COVID-19.

There were 6 and 4 variants associated with circulating ADAM17 at extracellular and cytoplasmic domain, respectively (LD r2<0.001, p < 5 x 10^−8^ for extracellular domain; p < 1 x 10^−6^ for cytoplasmic domain). These variants were used as instrument variables to examine the effect of circulating ADAM17 on COVID-19 with average F-statistic of 90 and 30 for extracellular and cytoplasmic domains, respectively (Table 1).

**Table 1.** Characteristics of single nucleotide polymorphisms used as instrument variables for circulating ADAM17 in primary Mendelian randomization analyses.

| Exposure | SNP | Chr | Pos | A1 | A2 | EAF | B | SE | P | N | F statistic |
| --- | --- | --- | --- | --- | --- | --- | --- | --- | --- | --- | --- |
| ADAM17<br>(extracellular) | rs10922098 | 1 | 196 664 651 | T | C | 0.599 | -0.129 | 0.008 | 6.3E-57 | 35338 | 252.8 |
|  | rs7549171 | 1 | 197 177 632 | G | A | 0.242 | 0.083 | 0.009 | 2.9E-19 | 35338 | 80.5 |
|  | rs1355538 | 3 | 165 505 177 | G | A | 0.602 | -0.066 | 0.008 | 3.1E-16 | 35338 | 66.8 |
|  | rs6457457 | 6 | 31 878 108 | T | C | 0.065 | 0.102 | 0.016 | 2.6E-10 | 35441 | 39.9 |
|  | rs444921 | 6 | 31 932 177 | T | C | 0.139 | 0.091 | 0.011 | 2.3E-15 | 35441 | 62.8 |
|  | rs28688825 | 6 | 32 587 157 | G | A | 0.196 | 0.061 | 0.010 | 1.2E-09 | 35439 | 37.0 |
| ADAM17<br>(cytoplasmic) | rs374896 | 1 | 196 692 378 | C | T | 0.568 | -0.040 | 0.008 | 3.1E-07 | 35339 | 26.2 |
|  | rs17209907 | 6 | 32 446 261 | T | C | 0.386 | 0.055 | 0.008 | 1.5E-11 | 35439 | 45.6 |
|  | rs12156434 | 9 | 124 133 218 | C | T | 0.101 | -0.066 | 0.013 | 5.2E-07 | 35374 | 25.2 |
|  | rs55701306 | 17 | 16 842 447 | T | C | 0.048 | 0.092 | 0.018 | 7.3E-07 | 35357 | 24.5 |

SNP: single nucleotide polymorphism; Chr: chromosome; Pos: position A1: effect allele; A2: other allele; EAF: effect allele frequency; B: beta; SE: standard error; P: p-value; N: sample size

Primary MR analysis (Figure 1A, Supplemental Table 2) showed that 1SD increase of genetically determined circulating ADAM17 (extracellular domain) were associated with increasing risk of developing critical COVID-19 (odds ratio [OR]=1.26, 95% CI 1.03-1.55, IVW). This potential causal effect was also observed in sensitivity MR analysis such as the weighted median and RAPS, but confidence intervals became broader in MR-Egger, possibly due to a less value of 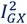 statistics was observed suggesting “no measurement error” assumption may be violated. No heterogeneity in the SNP effects was found, nor directional horizontal pleiotropy was detected (Table 2). Circulating ADAM17 (extracellular domain) showed a trend in the risk of the hospitalized COVID-19 (OR=1.09, 95% CI 0.99-1.21) but not for SARS-CoV-2 infection. Casual effects for the cytoplasmic domain of ADAM17 on COVID-19 were not observed.

**Figure 1.**
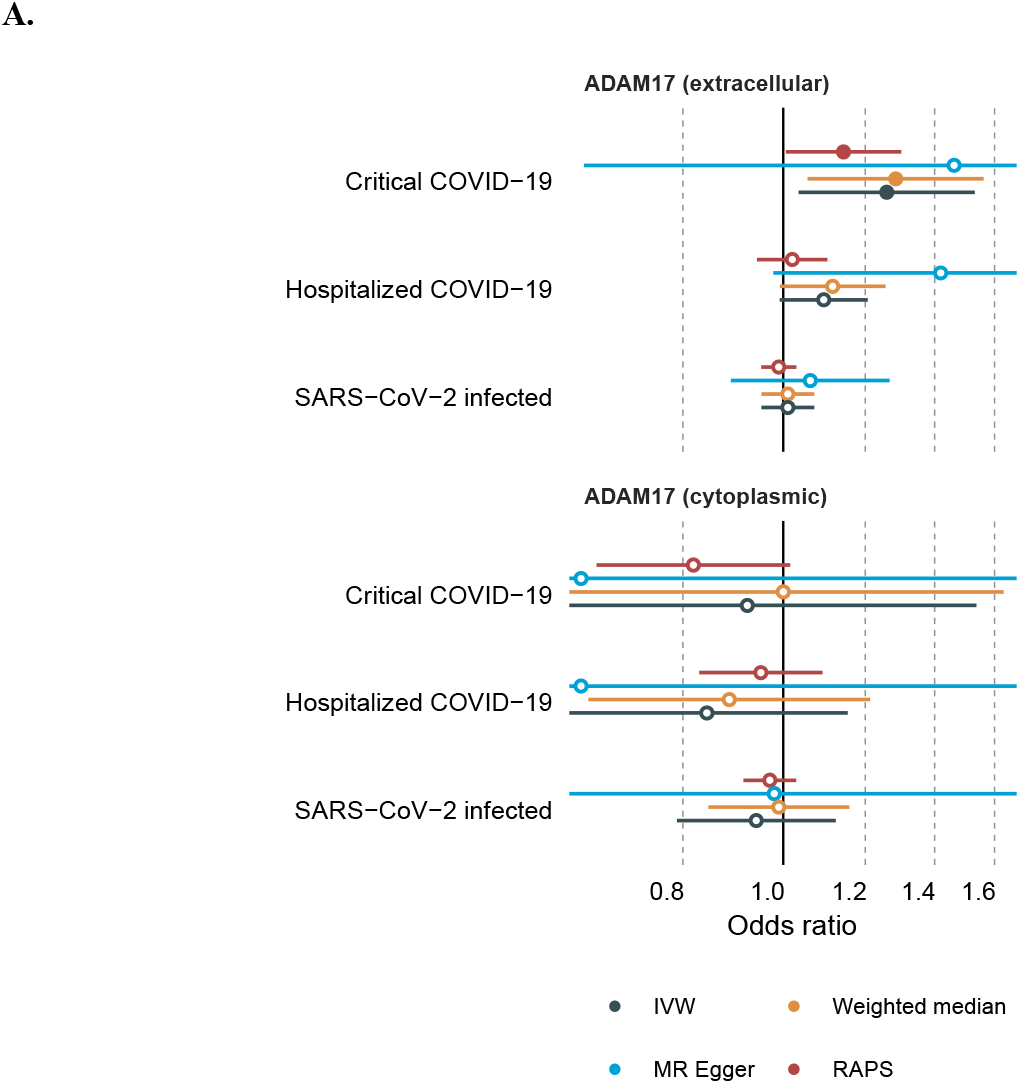
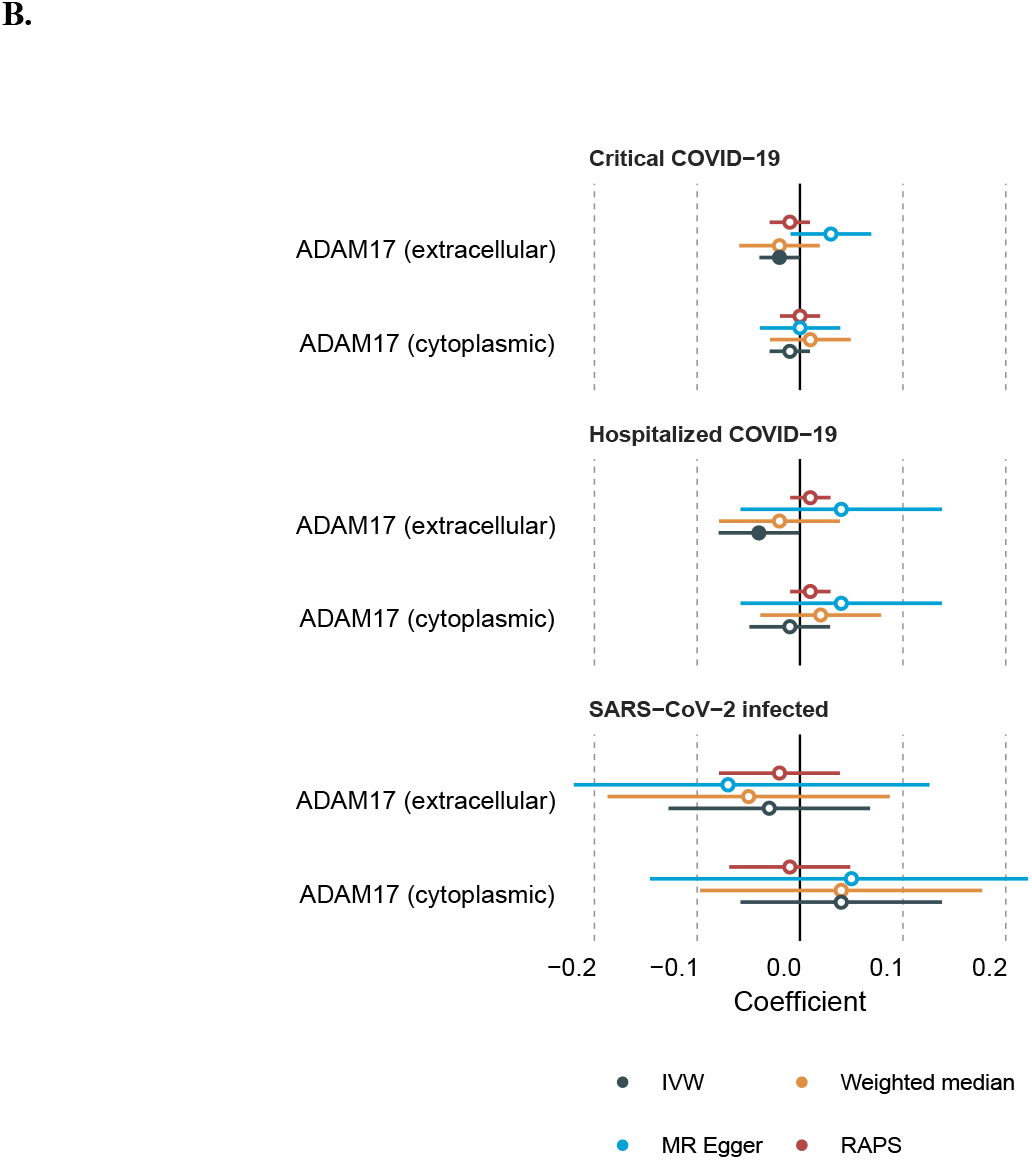
Forest plots showing results from Mendelian randomization analyses. (A) Causal effect for circulating ADAM17 on COVID-19. (B) Causal effect for COVID-19 on circulating ADAM17. IVW: inverse-variance weighted; RAPS: robust adjusted profile score

**Table 2.** Heterogeneity and directional horizontal pleiotropy.

| Exposure | Outcome | Heterogeneity |  | Horizontal pleiotropy |  |  |
| --- | --- | --- | --- | --- | --- | --- |
|  |  | Cochran's Q | P | Egger intercept | SE | P |
| ADAM17<br>(extracellular) | Critical COVID-19 | 9.14 | 0.10 | -0.014 | 0.039 | 0.74 |
|  | Hospitalized COVID-19 | 4.42 | 0.49 | -0.025 | 0.018 | 0.24 |
|  | SARS-CoV-2 infected | 3.18 | 0.67 | -0.005 | 0.009 | 0.61 |
| ADAM17<br>(cytoplasmic) | Critical COVID-19 | 2.97 | 0.23 | 0.023 | 0.075 | 0.81 |
|  | Hospitalized COVID-19 | 2.72 | 0.26 | 0.017 | 0.039 | 0.74 |
|  | SARS-CoV-2 infected | 8.15 | 0.04 | -0.002 | 0.023 | 0.93 |

P: p-value; Egger intercept: MR-Egger intercept ; SE: standard error;

Multivariable MR analysis suggested a direct causal role of ADAM17 (extracellular domain) on critical COVID-19 (OR=1.09; 95% CI 1.01-1.17), accounting for BMI, a risk fact suggested being causal in severe COVID-19 (17, 18).

Reversely, causal effects for COVI-19 on ADAM17 were not observed in general. Critical COVID-19 seems to have a marginal negative effect on circulating ADAM17 (extracellular domain, OR=0.98, 95%CI 0.95-1.00, IVW), so did hospitalized COVID-19 on circulating ADAM17 (extracellular domain, OR=0.96, 95%CI 0.92-1.00, IVW) (Figure 1B, Supplemental Table 3). However, these cannot be further confirmed in sensitivity MR analyses.

## Discussion

In the present study, we observe evidence for a causal effect of circulating ADAM17 (extracellular domain) on severe COVID-19. These findings add elegantly to recent studies showing that increasing levels of ADAM17 substrates (ACE2, TNFR1 and TNFR2) are associated with adverse clinical outcome in COVID-19 patients (6), and that inhibition of ADAM17 activity *in vitro* inhibits SARS-CoV-2 infection dose-dependently (19). Together, these observations suggest that the attenuation of ADAM17 activity could potentially mitigate the severity of critical COVID-19 cases.

Previous studies showed circulating levels of the extracellular domain of ADAM17 were higher in individuals with metabolic syndrome, type 2 diabetes, obesity, and correlated positively with several known cardiovascular risk markers including blood pressure, triglycerides, cholesterol and C-reactive protein (9). Additionally, genetically explained body-mass index and smoking was causally related to the risk of risk of being hospitalized with COVID-19 (10). High levels of circulating GCNT4, RAB14, C1GALT1C1, CD207 and ABO were also suggested being causally associated with an increased risk of critical COVID-19 with comparable odds ratios varying from 1.12 to 1.35 (20).

A pre-existing cardiometabolic dysfunction is associated with endothelial injury, ongoing inflammation and increased ADAM17 activity (8). Based on the present study, it could be speculated that a greater pre-existing ADAM17 expression, once infected by SARS-CoV-2, leads to increased shedding of ADAM17 substrates, including ACE2 and TNF-alpha. This could lead to exacerbation of the pre-existing endothelial dysfunction, as well as a further dysregulation of the renin-angiotensin system and immune system in these patients, which could lead to severe disease.

An important limitation of the study is that circulating ADAM17 does not necessarily reflect the ADAM17 activity in the cells/tissue. Also, other sheddases may shed ACE2 and inflammatory mediators. However, under pro-inflammatory stimuli, the sheddase activity by ADAM17 is favored over, at least, ADAM10, promoted by increased iRhom2 expression (3). In what concerns the discrepancy between the extracellular and cytoplasmic domains of ADAM17 and COVID-19 severity, only speculation is possible currently. To the best of our current knowledge, it is understood that ADAM17 can undergo swift posttranslational activation primarily through its transmembrane domain, whereas its cytoplasmic domain does not contribute significantly to this activation process (21-23). Another explanation could be that part of the measured ADAM17 reflects ADAM17 bound to microparticles from endothelial cells, platelets and leukocytes, where the cytoplasmic domain is hidden from detection (19). Strengthening this hypothesis, strong positive correlations between the extracellular domain of ADAM17, and platelet and white blood cell counts were shown (9). There might be potential overlap between samples from the GWAS of ADAM17 and COVID-19. However, the bias caused by this (if any) is likely negligible as estimated by the previously published method (https://sb452.shinyapps.io/overlap) (24). It should be also noted that Mendelian randomization may not fully rule out potential collider bias or selection bias (25) even when multiple Mendelian randomization method and GWAS with largest possible sample sizes were employed in the present study. Further studies are needed to ascertain the causal role of ADAM17 in the risk of severe COVID-19. Indeed, protective effect of ADAM17 inhibition was also suggested using mouse model (7).

In conclusion, genetic susceptibility to the increased circulating ADAM17 (extracellular domain) is associated with a high risk of critical COVID-19. In conjunction with prior research, the timely selective inhibition of ADAM17 emerges as a potential therapeutic avenue worthy of investigation against severe COVID-19.

## Data Availability

All data produced in the present study are available upon reasonable request to the authors

https://www.decode.com/summarydata/

https://www.ebi.ac.uk/gwas/studies/GCST011074

https://mrcieu.github.io/TwoSampleMR/articles/introduction.html

