## Supplemental Table 1-3 for "Circulating ADAM17 is associated with COVID-19 severity"

**Supplemental Table 1. Genome-wide association summary statistics used in the MR analyses.**

| Trait | Name of the dataset | Sample size | Consortium or project name | PMID | link |
| --- | --- | --- | --- | --- | --- |
| ADAM17 (extracellular) | 8959_61_ADAM17 | 35 559 | Icelandic Cancer Project and deCODE | 34857953 | https://www.decode.com/summarydata/ |
| ADAM17  (cytoplasmic) | 8821_1_ADAM17 | 35 559 | Icelandic Cancer Project and deCODE | 34857953 | https://www.decode.com/summarydata/ |
| Critical COVID-19 | A2_ALL_eur_leave_23andme | 1 086 211 | COVID-19 host genetics initiative | 34237774 | https://www.covid19hg.org/results/r7/ |
| Hospitalized COVID-19 | B2_ALL_eur_leave_23andme | 2 095 324 | COVID-19 host genetics initiative | 34237774 | https://www.covid19hg.org/results/r7/ |
| SARS-CoV-2 infected | C2_ALL_eur_leave_23andme | 2 597 856 | COVID-19 host genetics initiative | 34237774 | https://www.covid19hg.org/results/r7/ |
| BMI | bmi.giant-ukbb.meta-analysis.combined | 806 834 | UKB and GIANT | 30239722 | https://portals.broadinstitute.org/collaboration/giant/images/1/14/Bmi.giant-ukbb.meta-analysis.combined.23May2018.HapMap2_only.txt.gz |

**Supplemental Table 2. Results from MR analysis estimating causal effects for circulating ADAM17 on COVID-19.**

| Exposure | Outcome | Method | #SNPs | beta | se | pval | mean  F statistic | I^2^ (unweighted) |
| --- | --- | --- | --- | --- | --- | --- | --- | --- |
| ADAM17  (extracellular) | Critical COVID-19 | MR Egger | 6 | 0.38 | 0.42 | 0.419 | 89.98 | 0.88 |
|  |  | IVW | 6 | 0.23 | 0.10 | 0.025 |  |  |
|  |  | Weighted median | 6 | 0.25 | 0.10 | 0.016 |  |  |
|  |  | RAPS | 23 | 0.13 | 0.07 | 0.041 | 39.76 | 0.84 |
|  | Hospitalized COVID-19 | MR Egger | 6 | 0.35 | 0.19 | 0.146 | 89.98 | 0.88 |
|  |  | IVW | 6 | 0.09 | 0.05 | 0.090 |  |  |
|  |  | Weighted median | 6 | 0.11 | 0.06 | 0.079 |  |  |
|  |  | RAPS | 23 | 0.02 | 0.04 | 0.614 | 40.20 | 0.84 |
|  | SARS-CoV-2 infected | MR Egger | 6 | 0.06 | 0.09 | 0.531 | 89.98 | 0.88 |
|  |  | IVW | 6 | 0.01 | 0.03 | 0.584 |  |  |
|  |  | Weighted median | 6 | 0.01 | 0.03 | 0.723 |  |  |
|  |  | RAPS | 22 | -0.01 | 0.02 | 0.719 | 40.76 | 0.85 |
| ADAM17  (cytoplasmic) | Critical COVID-19 | MR Egger | 3^a^ | -0.45 | 1.27 | 0.782 | 25.30 | 0.75 |
|  |  | IVW | 3^a^ | -0.08 | 0.26 | 0.758 |  |  |
|  |  | Weighted median | 3^a^ | 0.00 | 0.25 | 0.993 |  |  |
|  |  | RAPS | 17 | -0.20 | 0.11 | 0.055 | 23.41 | 0.69 |
|  | Hospitalized COVID-19 | MR Egger | 3^a^ | -0.45 | 0.67 | 0.626 | 25.30 | 0.75 |
|  |  | IVW | 3^a^ | -0.17 | 0.16 | 0.283 |  |  |
|  |  | Weighted median | 3^a^ | -0.12 | 0.16 | 0.466 |  |  |
|  |  | RAPS | 17 | -0.05 | 0.07 | 0.467 | 23.33 | 0.70 |
|  | SARS-CoV-2 infected | MR Egger | 4 | -0.02 | 0.40 | 0.966 | 30.36 | 0.62 |
|  |  | IVW | 4 | -0.06 | 0.09 | 0.524 |  |  |
|  |  | Weighted median | 4 | -0.01 | 0.08 | 0.900 |  |  |
|  |  | RAPS | 17 | -0.03 | 0.03 | 0.304 | 23.38 | 0.70 |

^a^ rs17209907 is not present in the outcome GWAS. IVW: Inverse-variance weighted; RAPS: robust adjusted profile score; I^2^: an adapted I-squared statistic to assess violation of the “NO Measurement Error” (NOME) assumption for instruments used for MR-Egger.

regression.

**Supplemental Table 3. Results from MR analysis estimating causal effects for COVID-19 on circulating ADAM17.**

| Exposure | Outcome | Method | #SNPs | beta | se | pval | mean  F statistic | I^2^ (unweighted) |
| --- | --- | --- | --- | --- | --- | --- | --- | --- |
| Critical COVID-19 | ADAM17  (extracellular) | MR Egger | 37 | 0.03 | 0.02 | 0.278 | 67.15 | 0.93 |
|  |  | IVW | 37 | -0.02 | 0.01 | 0.034 |  |  |
|  |  | Weighted median | 37 | -0.02 | 0.02 | 0.199 |  |  |
|  |  | RAPS | 107 | -0.01 | 0.01 | 0.342 | 38.06 | 0.88 |
| Hospitalized COVID-19 |  | MR Egger | 40 | 0.04 | 0.05 | 0.461 | 51.25 | 0.85 |
|  |  | IVW | 40 | -0.04 | 0.02 | 0.048 |  |  |
|  |  | Weighted median | 40 | -0.02 | 0.03 | 0.362 |  |  |
|  |  | RAPS | 141 | 0.01 | 0.01 | 0.572 | 30.48 | 0.77 |
| SARS-CoV-2 infected |  | MR Egger | 16 | -0.07 | 0.10 | 0.472 | 72.00 | 0.94 |
|  |  | IVW | 16 | -0.03 | 0.05 | 0.486 |  |  |
|  |  | Weighted median | 16 | -0.05 | 0.07 | 0.424 |  |  |
|  |  | RAPS | 78 | -0.02 | 0.03 | 0.553 | 33.17 | 0.86 |
| Critical COVID-19 | ADAM17  (cytoplasmic) | MR Egger | 37 | 0.00 | 0.02 | 0.90 | 67.15 | 0.93 |
|  |  | IVW | 37 | -0.01 | 0.01 | 0.19 |  |  |
|  |  | Weighted median | 37 | 0.01 | 0.02 | 0.76 |  |  |
|  |  | RAPS | 107 | 0.00 | 0.01 | 0.63 | 38.06 | 0.88 |
| Hospitalized COVID-19 |  | MR Egger | 40 | 0.04 | 0.05 | 0.43 | 51.25 | 0.85 |
|  |  | IVW | 40 | -0.01 | 0.02 | 0.75 |  |  |
|  |  | Weighted median | 40 | 0.02 | 0.03 | 0.38 |  |  |
|  |  | RAPS | 141 | 0.01 | 0.01 | 0.32 | 30.48 | 0.77 |
| SARS-CoV-2 infected |  | MR Egger | 16 | 0.05 | 0.10 | 0.60 | 72.00 | 0.94 |
|  |  | IVW | 16 | 0.04 | 0.05 | 0.45 |  |  |
|  |  | Weighted median | 16 | 0.04 | 0.07 | 0.53 |  |  |
|  |  | RAPS | 78 | -0.01 | 0.03 | 0.73 | 33.17 | 0.86 |

IVW: Inverse-variance weighted; RAPS: robust adjusted profile score; I^2^: an adapted I-squared statistic to assess violation of the “NO Measurement Error” (NOME) assumption for instruments used for MR-Egger.

regression.
